# Demonstration of antibodies against SARS-CoV-2, neutralizing or binding, in seroconversion panels after mRNA-1273, BNT-162b2 and Ad26.COV2.S vaccine administration

**DOI:** 10.1101/2022.03.28.22272552

**Authors:** Francisco Belda, Oscar Mora, Monica Lopez-Martinez, Nerea Torres, Ana Vivanco, Silvia Marfil, Edwards Pradenas, Marta Massanella, Julià Blanco, Rebecca Christie, Michael Crowley

## Abstract

Seroconversion panels were collected before and after vaccination with three COVID-19 vaccines: two mRNA vaccines (mRNA-1273 and BNT-162b2) and one adenovirus vector vaccine (Ad26.COV2.S). The panels were tested for antibody activity by chemiluminescent immunoassay, ELISA and one was tested in a pseudovirus neutralization assay. Participants positive for anti-SARS-CoV-2 antibodies before vaccination (18.6%) had a higher response to the first vaccine dose than participants who tested negative. For two-dose vaccines, older participants showed a lower response to the first dose than younger participants. All participants showed positive antibody responses after the second vaccine. For the adenovirus vector vaccine, two participants did not generate antibody responses two weeks and two months after vaccination. Three participants were negative at two weeks but positive at two months. Pseudovirus neutralization showed good correlation with antibody activity (correlation coefficient =0.78, p<0.0001). Antibody responses in participants over 45 years old tended to be less robust.

## Introduction

Since the start of the SARS-CoV-2 pandemic, nearly 30 pharmaceutical companies, research institutes and academic laboratories worldwide have been working toward the development of effective vaccines against COVID-19. These efforts have taken various approaches including mRNA, DNA, protein subunits, and attenuated or non-replicating viruses. [1] Among the vaccines utilizing mRNA and nanotechnology, mRNA-1273 (Moderna) and BNT-162b2 (Pfizer/BioNTech) are the most advanced.

Within one year of the emergence of the SARS-CoV-2 virus, these two novel and effective mRNA vaccines became available under emergency use authorization (EUA) by the US Food and Drug Administration (FDA). The BNT162b2 vaccine contains 30 μg of SARS-CoV-2 full-length spike (pre-fusion conformation) mRNA administered as two doses three weeks apart. The mRNA-1273 vaccine 100 μg of pre-fusion-stabilized spike glycoprotein mRNA given as two doses four weeks apart. [2]

The BNT-162B2 vaccine (Comirnaty®, BioNTech, Mainz, Germany/Pfizer, New York, NY, USA) was first utilized under an EUA beginning December 11, 2020 [3] and became the first COVID-19 vaccine approved by the FDA on August 23, 2021. It was approved for the prevention of COVID-19 in individuals 16 years of age and older. In addition, the BNT-162B2 vaccine is available for use under an emergency use authorization (EUA) for children ages 12 to 15 years and as a third booster dose in immunocompromised individuals, people over 65, individuals between 16 and 64 who are at high risk of severe COVID-19 and person with high environmental exposure to SARS-CoV-2 which puts them at risk for severe COVID-19. [4]

The FDA issued an EUA for the mRNA-1273 vaccine for the prevention of COVID-19 in individuals over 18 years of age on December 18, 2020. As with the EUA for BNT-162b2, the EUA for mRNA-1273 was amended in August 2021 to allow the administration of an additional dose to certain immunocompromised individuals. [5]

The Ad26.COV2.S vaccine (Janssen/Johnson & Johnson, Beerse, Belgium) uses a recombinant, replication-incompetent human adenovirus type 26 vector. This vector encodes for a full SARS-CoV-2 spike protein in a stabilized pre-fusion conformation. A single dose of Ad26.COV2.S protected vaccinated individuals against symptomatic and asymptomatic SARS-CoV-2 infection. For those who did get infected, the vaccine was effective in preventing severe disease including hospitalization and death. [6]

Infection with SARS-CoV-2 or vaccination with an effective vaccine initiates an immune response with includes the production of antibodies that bind to viral proteins, i.e., binding antibodies.[7] Not all binding antibodies can block cellular infiltration and/or replication by the SARS-CoV-2 virus. The subpopulation of antibodies that can block these processes are called neutralizing antibodies (NAbs). It is unknown when in the course of infection or after vaccination NAbs are produced or if they are produced from the onset of antibody formation. While most individuals produce binding antibodies in response to SARS-CoV-2 infection, not all will develop NAbs to SARS-CoV-2.[8]

Most SARS-CoV-2 infections induce a response in the adaptive immune system. [9] NAbs that prevent the virus from binding to the host-cell receptor, angiotensin converting enzyme 2 (ACE2) are essential for this immune response. NAbs target the receptor binding domain (RBD) of the viral spike protein and thereby inhibit ACE2 binding. [10, 11] These NAbs may be of therapeutic value through treatments such as convalescent plasma [12], and hyperimmune globulins. [13] In addition, they can serve as structural templates for vaccine development. Consequently, there is a need for assays that reliably and efficiently identify the most potent NAbs. The gold standard to detect NAbs is a virus neutralization test conducted on live cells which can be performed with live virus or psuedoviruses. [14] The level of neutralizing antibodies that confers immunity to infection/reinfection with SARS-CoV-2 has yet to be elucidated. [15]

Seroconversion panels are a series of blood samples collected before and after the development of antibodies in response to viral infection or vaccination. These panels can be useful in the development of antibody assays, determination of the window period of detection (the period between infection and the appearance of antibodies), and the development (validation) and manufacture (quality control) of commercial antibody tests. Reliable detection of viral exposure and vaccine effectiveness is a critical step in gaining control of the current COVID-19 pandemic and allowing the general population to return to normal ways of life. The panels can also be a source of well-defined neutralizing antibodies (before and after vaccination) that can be useful in investigating their effectiveness against new SARS-CoV-2 variants. The studies in this paper describe the characterization of seroconversion panels from donors vaccinated for COVID-19 with the three vaccines currently available in the US: BNT-162b2 (Pfizer/BioNTech), mRNA-1273 (Moderna) and Ad26.COV2.S (Janssen/Johnson & Johnson).

## Materials and Methods

### Seroconversion Panel Collection

The seroconversion panels described in this paper were derived from samples collected at a hospital from volunteer donors. The donors provided informed consent and their samples were collected under an approved IRB protocol ([1149706-4] Diagnostic QC and Pre-Clinical Sample Collection Project: Ballad Health System Institutional Review Board (IRB #00003204), Johnson City, TN, USA). These studies were conducted in compliance with all applicable regulatory guidelines. The seroconversion panels were comprised of serum samples collected before and after administration of the COVID-19 vaccines. This study characterizes the appearance of anti-SARS-CoV-2 antibodies after vaccine administration.

Four seroconversion panels were analyzed in this study: one mRNA-1273 (Moderna) n = 15 donors; two BNT-162b2 (Pfizer/BioNTech) n = 15 donors for each panel; and one Ad26.COV2.S (Janssen/Johnson & Johnson) n= 14 donors. For mRNA-based vaccines (mRNA-1273 and BNT-162b2) panels, samples were collected prior to the first vaccination (objective target ≤ 2 days: sample 1), prior to the second vaccination (objective target ≤ 2 days: sample 2), and after the second vaccination (objective target 13-15 days; sample 3). For Ad26.COV2.S, samples were collected on vaccination day (prior to the injection: sample 1), two weeks after vaccination (14 days: sample 2) and 2 months after vaccination (59-62 days: sample 3). Samples were collected as plasma (in the presence of potassium EDTA) and/ or serum (in serum separating tubes).

As previously described [16], the mRNA-1273 panel was made up of undiluted, unpreserved serum samples collected from 15 participants between 22 December 2020 and 25 February 2021. The donors were health adults (21-76 years old) who received the prescribed course of mRNA-1273 vaccines (two injections of 100 µg -objective target 28 days apart). There were 10 female and 5 male donors, and all were white/Caucasian.

The two BNT-162b2 panels were collected at different times. The first panel (BNT-162b2 Panel 1) was collected between 19 February 2021 and 23 April 2021. The second panel (BNT-162b2 Panel 2) was collected between 25 May 2021 and 24 September 2021. The BNT-162b2 panels were either undiluted, unpreserved serum or EDTA-treated plasma collected from 15 donors each. These donors were healthy adults 36-70 years old for the first panel and 29 to 67 years old for the second panel. The participants for both panels received the approved vaccination regimen (two injections of BNT-162b2 30 µg – objective target 21 days apart). There were 7 female and 8 male donors in panel 1 and 9 female and 6 male donors in panel 2. All the donors were white/Caucasian.

The Ad26.COV2.S panel was undiluted, unpreserved serum collected from 14 donors between 7 March 2021 and 24 September 2021. These donors were between 31 and 64 years of age. There were equal numbers (n=7) of female and male donors and all were white/Caucasian. These donors received the approved regimen of a single injection (0.5 mL) of the Ad26.COV2.S vaccine.

All samples were stored at -20°C until analyzed. Prior to analysis, samples were thawed at room temperature and mixed by inversion.

### CLIA and ELISA Assays

The samples in the seroconversion panels were tested for the presence of anti-SARS-CoV-2 antibodies using chemiluminescent immunoassays (CLIA: Liaison SARS-CoV-2 IgG assay, Diasorin, Inc., Saluggia, Italy; EUA approved) and enzyme-linked immunosorbent assays (ELISA: Progenika anti-SARS-CoV-2 IgG Kit, Progenika Biopharma, Derio, Bizkaia, Spain; CE-IVD certified immunoassay). These assays were performed following the directions provided by the manufacturers. The Diasorin assay utilizes antigens to the S1 and S2 spike protein IgG while the Progenika assay utilizes only antigens to S1 IgG. The specific sequences antigens are not disclosed. Since the assays were developed independently, they are assumed to be non-identical.

A kappa correlation (JMP software 16.0) statistic test was performed to compare the qualitative values between CLIA and ELISA assays.

### Neutralizing Antibody Determination

#### Cell line

HEK293T cells (presumably of female origin) overexpressing wild-type human ACE-2 (Integral Molecular, USA) were used as target for SARS-CoV-2 spike expressing pseudovirus infection. Cells were maintained in T75 flasks with Dulbecco′s Modified Eagle′s Medium (DMEM) supplemented with 10% fetal bovine serum and 1μg/mL puromycin).

#### Pseudovirus generation

Pseudoviruses (HIV reporter) expressing the SARS-CoV-2 S protein and luciferase were constructed using the defective HIV plasmid, pNL4-3.Luc.R-.E-. The plasmid was obtained from the NIH AIDS Reagent Program. [17] Expi293F cells were transfected using ExpiFectamine293 Reagent (Thermo Fisher Scientific, Waltham, MA, USA) with pNL4-3.Luc.R-.E-and SARS-CoV-2.SctΔ19 (Wuhan, G614 or B.1.1.7); at an 8:1 ratio, respectively. Control pseudoviruses were generated by replacing the S protein expression plasmid with a vesicular stomatitis virus (VSV)-G protein expression plasmid as previously reported. [18] Supernatants were harvested 48 hours after transfection, filtered at 0.45 μm, frozen, and titrated on HEK293T cells overexpressing wild-type human ACE-2.

### Neutralization assays

Neutralization assays were performed in duplicate. Briefly, 200X the median tissue culture infectious dose (TCID50) of pseudovirus was preincubated with three-fold serial dilutions (1/60–1/14,580) of heat-inactivated plasma samples in Nunc 96-well cell culture plates (Thermo Fisher Scientific) for 1 h at 37°C. Then 2 × 10^4^ HEK293T/hACE2 cells treated with DEAE-Dextran (Sigma-Aldrich, St. Louis, MO, USA) were added. Results were read after 48 h using an EnSight Multimode Plate Reader and BriteLite Plus Luciferase reagent (Perkin Elmer, Waltham, MA, USA).

The results were normalized and the ID50 (the reciprocal dilution inhibiting 50% of the infection) was calculated by plotting the log of plasma dilution versus response and fitting to a 4-parameter equation in Prism 8.4.3 (GraphPad Software, San Diego, CA, USA). This neutralization assay has been previously validated in a large subset of samples. [7]

## RESULTS

### mRNA-1273 (Moderna) seroconversion panel

As previously described [16], the pre-vaccination samples in the mRNA-1273 panel showed positive values for two participants suggesting that they had been previously infected with SARS-CoV-2 (participants 5 and 15). The post-vaccination samples collected from these participants after the first dose showed the largest antibody responses in the panel. After the first dose of mRNA-1273, samples from 13 of the 15 participants showed positive antibody response by CLIA and 14 of 15 by ELISA. The two participants who tested negative after the first dose of vaccine were the oldest participants in the panel (> 70 years old: participants 3 and 4). After the second dose, samples for all 15 of the participants were positive for anti-SARS-CoV-2 antibodies (in both assays). Overall, there was good agreement between the results of the CLIA and ELISA assays (Kappa coefficient = 0.947). The value of the equivocal range was eliminated to perform this statistical test.

The post-vaccination second dose samples from the mRNA-1273 panel were tested for neutralizing antibodies. The samples from all participants were positive for neutralizing antibodies (Figure 1). When the neutralizing antibody results were compared with the ELISA antibody detection results, a positive correlation was observed in both analyses (Correlation coefficient=0.78 and p-value<0.0001).

**Figure 1.**
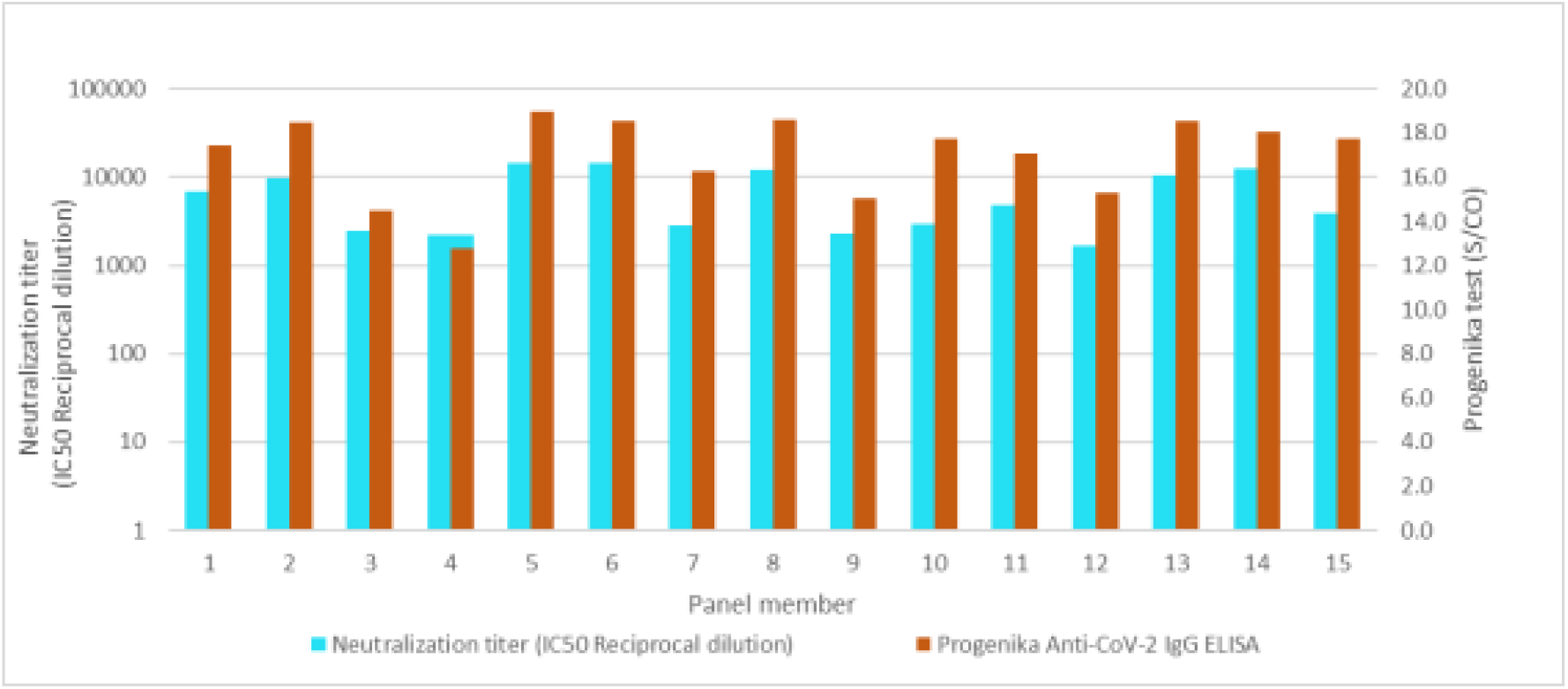
Antibody responses in a seroconversion panel of 15 participants after vaccination with two doses of the mRNA-1273 SARS-CoV-2 vaccine (Moderna). The column in blue, shows the results of neutralizing antibody titers expressed in (IC_50_ Reciprocal Dilution). Responses ≥ 250 were considered positive. The column in brown, shows the antibody responses obtained using an enzyme-linked immunosorbent assay (ELISA). Responses ≥ 1.1 S/CO were considered positive.

### BNT-162b2 (Pfizer/BioNTech) seroconversion panels

Testing (CLIA) of the pre-vaccination samples in BNT-162b2 Panel 1 gave the following results (Figure 2; first point): 12 negative and 3 positive results (participants 2, 5 and 15). The detection of anti-SARS-CoV-2 antibodies in these three participants indicates that they were infected with the virus prior to collection of the samples.

**Figure 2.**
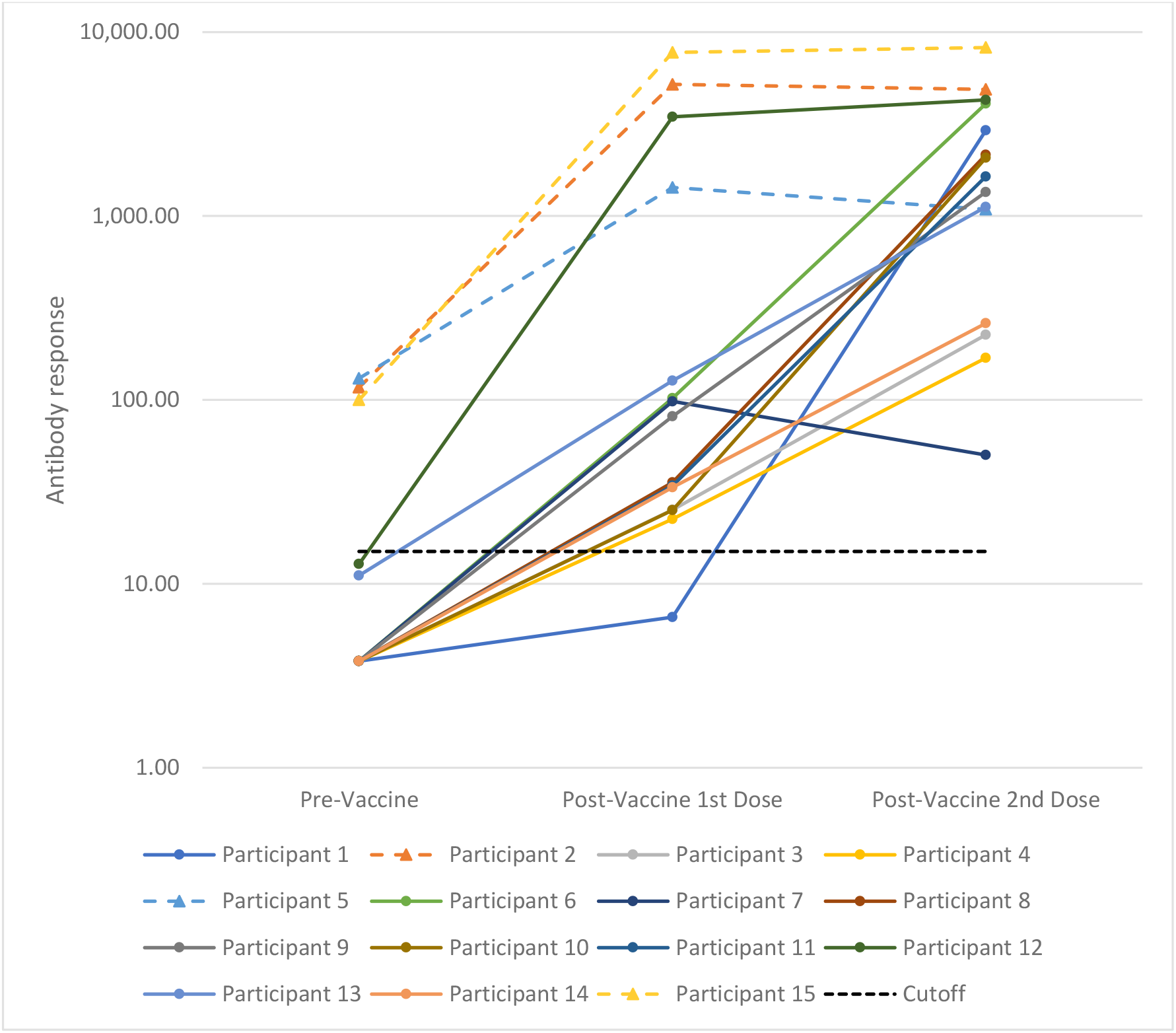
Antibody responses in a seroconversion panel of 15 subjects before and after vaccination with two doses of the BNT162b2 SARS-CoV-2 vaccine (Pfizer/BioNTech) -Panel 1. These results were obtained using a chemiluminescent immunoassay and are expressed as arbitrary units/mL (AU/mL). Responses ≥ 15.0 units were considered positive. The first point shows results from samples collected prior to the first vaccination. The second point shows the results from samples collected after the first vaccination and prior to the second vaccination. The third point shows the results from samples collected after the second vaccination. The dashed lines correspond to the convalescent participants, while the solid lines correspond to the naïve participants.

Sample collected after the first vaccine injection showed 14 positive and 1 negative results (Figure 2; second point). The largest antibody responses were detected in the three participants (2, 5 and 15) who had detectable antibodies in their pre-vaccination samples. The sample from participant 12 also showed a high antibody response to the first vaccine injection. The pre-vaccination sample from this participant was close to the cutoff value for positivity. The single negative sample collected after the first vaccine dose was in one of the older participants (participant 1; 55-60 years old).

Samples collected after the second vaccine injection were positive in all 15 participants (Figure 2; third point). Lower antibody responses were seen in four participants (3, 4, 7 and 14) compared to the rest of the participants. These participants were between 55 and 80 years old. The sample from participant 7 (60-65 years old) showed a lower level of antibody activity after the second vaccine dose that that seen after the first vaccine dose.

Results from CLIA testing of a second BNT-162b2 seroconversion panel (BNT-162b2 Panel 2) showed ten negative and 5 positive anti-SARS-CoV-2 antibody responses in the pre-vaccination samples (Figure 3; first point – participants 4, 8, 9, 10 and 15). As previously noted, the presence of anti-SARS-CoV-2 IgG in pre-vaccination samples indicates infection with SARS-CoV-2 prior to collection of the samples.

**Figure 3.**
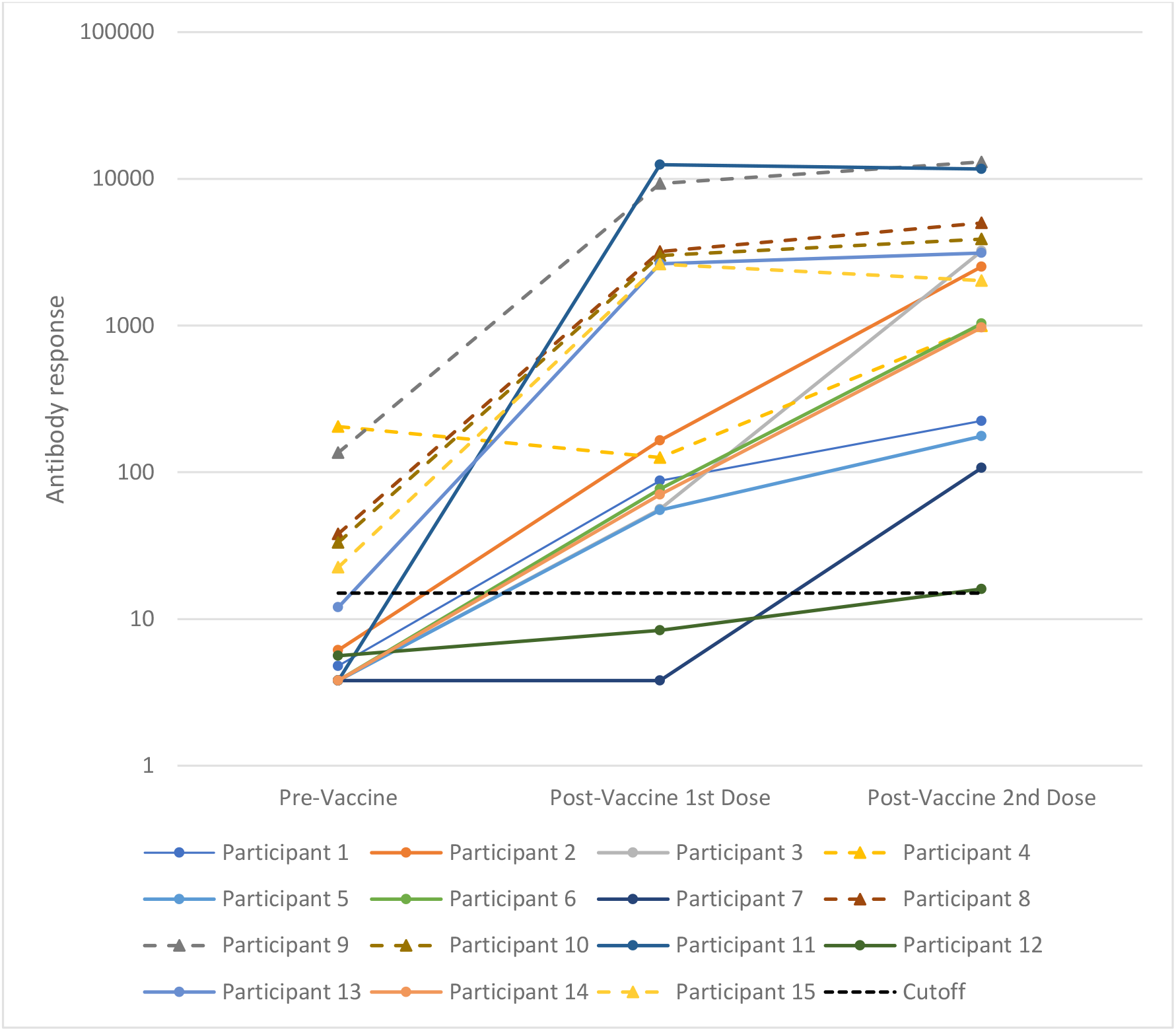
Antibody responses in a seroconversion panel of 15 subjects before and after vaccination with two doses of the BNT162b2 SARS-CoV-2 vaccine (Pfizer/BioNTech) – Panel 2. These results were obtained using a chemiluminescent immunoassay and are expressed as arbitrary units/mL (AU/mL). Responses ≥ 15.0 units were considered positive. The first point shows results from samples collected prior to the first vaccination. The second point shows the results from samples collected after the first vaccination and prior to the second vaccination. The third point shows the results from samples collected after the second vaccination. The dashed lines correspond to the convalescent participants, while the solid lines correspond to the naïve participants.

The samples in this panel collected after the first injection of the BNT-162b2 vaccine showed 13 positive and 2 negative results (Figure 3; second point). As with Panel 1, the greatest antibody responses were detected in the five participants that had detectable anti-SARS-CoV-2 antibodies in their pre-vaccination samples (participants 4, 8, 9, 10 and 15). Two participants had negative antibody responses after the first vaccine injection (participants 7 and 12). These participants were 45-50 and 25-30 years old.

The sample collected after the second BNT-162b2 vaccine injection were positive for all 15 participants (Figure 3; third point). The lowest activity was seen in participant 12 (25-30 years old) 16 AU/mL just above the cutoff value of 15 AU/mL. This participant (12) was one of the two participants that tested negative after the first vaccination. The other participants with low antibody responses (1, 5 and 7) were 45-60 years old.

### Ad26.COV2.S (Janssen/Johnson & Johnson) seroconversion panel

Testing of the pre-vaccination samples for the Ad26.COV2.S panel showed 13 negative and 1 positive anti-SARS-CoV-2 antibody responses (Figure 4; first point). The one positive sample indicates a previous SARS-CoV-2 infection in this participant (11) prior to the collection of the sample.

**Figure 4:**
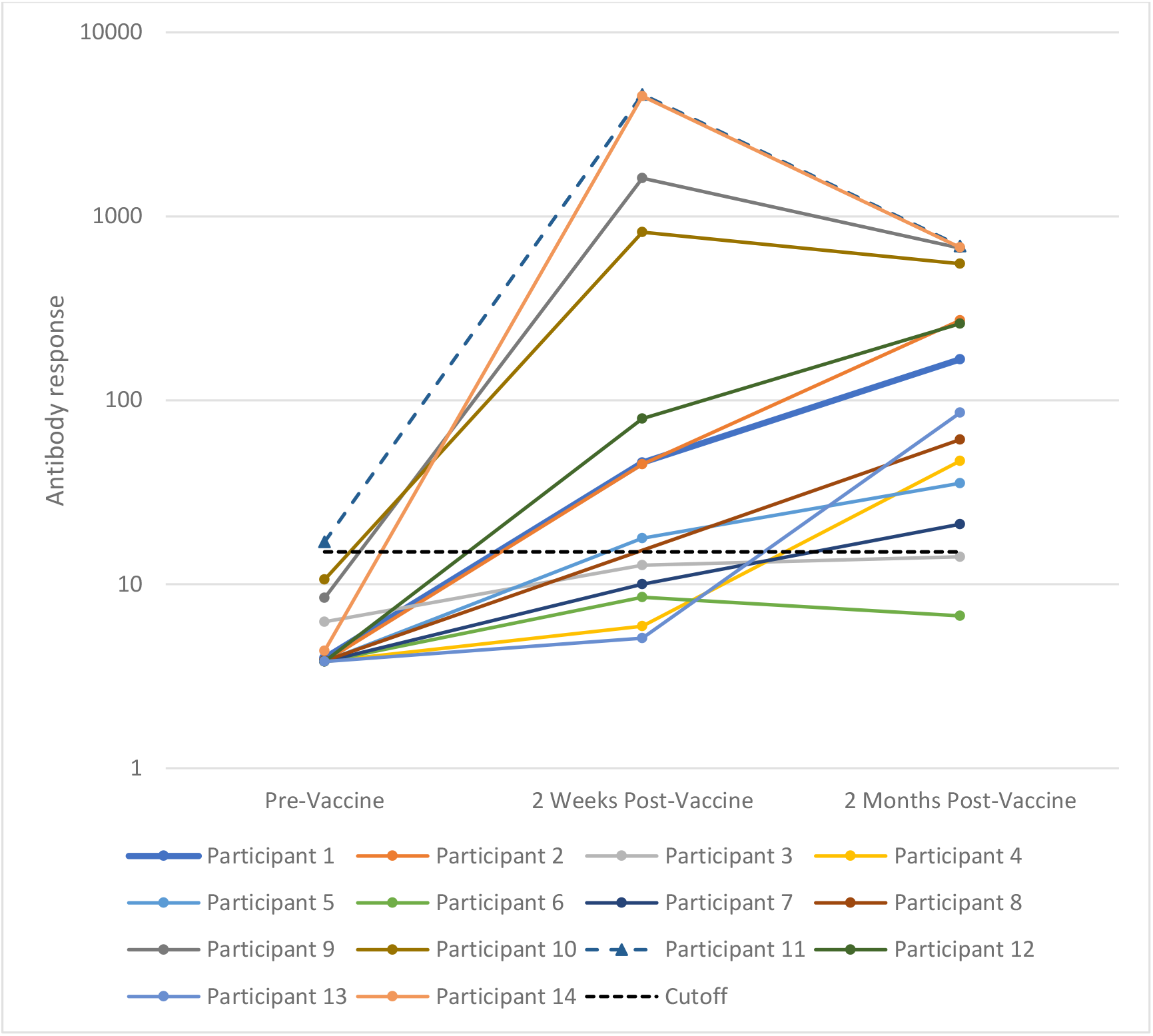
Antibody responses in a seroconversion panel of 14 subjects before vaccination with one dose of the Ad26.CoV2.S SARS-CoV-2 vaccine (Janssen/Johnson & Johnson). These results were obtained using a chemiluminescent immunoassay are expressed as arbitrary units/mL (AU/mL). Responses ≥ 15.0 units were considered positive. The first point shows results from samples collected prior the vaccination. The second point shows the results from samples collected two weeks after the vaccination. The third point shows the results from samples collected two months after vaccination The dashed lines correspond to the convalescent participants, while the solid lines correspond to the naïve participants.

The samples collected two weeks after the single-dose vaccination showed eight positive and 6 negative antibody responses (Figure 4; second point). Samples from 12 participants showed positive responses two months after vaccination (Figure 4; third point). Two participants (3 and 6) showed negative antibody responses after two weeks and two months. These participants were 40-45 and 60-65 years old. The three participants that showed a negative antibody response at two weeks but a positive response at two months were 30-35 (n=1) and 60-65 (n=2) years old.

## Discussion

Four seroconversion panels have been created by collecting serum or plasma samples from participants before and after COVID-19 vaccination. Three of the panels were drawn from participants who were administered one of the two-dose mRNA-based vaccines (mRNA-1273 – one panel and BNT-162b2 – two panels) and one panel from participants given a single dose of the replication-incompetent adenovirus-based vaccine (Ad26.COV2.S). These panels were tested using CLIA and ELISA technologies. Comparison of the CLIA and ELISA results for the mRNA-1273 panel showed good agreement between these assays. [16]

In addition, neutralization assays were conducted in this study and showed congruence between the neutralization assay and the CLIA. Neutralizing activity that blocks viral entry into cell has been shown to be responsible for antibody-mediated prevention of coronavirus infection. [19] Neutralizing anti-SARS-CoV-2 antibodies have been shown to mainly target the receptor-binding domains on the S1 region of the spike protein. [20] When immunoassay results correlate with neutralization assays, as in this study, the simpler, less labor-intensive immunoassays may be a useful initial indirect screen for antibody neutralization activity. A similar correlation between immunoassay and neutralization results has been seen in other studies. [21, 22]

The results from all the seroconversion panels illustrate several important points regarding antibody responses to vaccination against COVID-19: 1) There is a small but significant fraction (18.6%) of the general population (as measured by the participants in this study) that have been infected with SARS-CoV-2 and have antibodies to the virus prior to vaccination; 2) For the mRNA-based vaccines, the second dose raised anti-SARS-CoV-2 antibody levels in participants without a previous SARS-CoV-2 infection to the same levels as those participants who had been infected with SARS-CoV-2 and had antibodies prior to vaccination; 3) Antibody responses tended to be less robust in the participants over 45 years old.

## Data Availability

All data produced in the present work are contained in the manuscript.

## Acknowledgements

Michael K. James, Ph.D. is acknowledged for medical writing and Jordi Bozzo, Ph.D., CMPP for editorial assistance.

## Author Contributions

All authors have participated in the conception or design of the work, data collection and critical revision of the article. FB, OM, RC and NC conceived the study. FB, OM and MC provided administrative support. RC and MC provided study materials (developed the seroconversion panels). FB, OM, MLM, NT AV and RC participated in the collection and assembly of data. SF, EP, MM and JB performed the neutralization experiments. All authors participated in data analysis and interpretation, manuscript writing and gave approval of the final manuscript.

## References

1. Noor, R., Developmental Status of the Potential Vaccines for the Mitigation of the COVID-19 Pandemic and a Focus on the Effectiveness of the Pfizer-BioNTech and Moderna mRNA Vaccines. Curr Clin Microbiol Rep, 2021: p. 1–8.

2. Montoya, J.G., et al., Differences in IgG antibody responses following BNT162b2 and mRNA-1273 vaccines. bioRxiv, 2021.

3. Emergency Use Authorization (EUA) for an unapproved product - review memorandum: Pfizer-BioNTech COVID-19 Vaccine/BNT-162b2. 2020 12 Oct 2021]; Available from: https://www.fda.gov/media/144416/download.

4. Comirnaty and Pfizer-BioNTech COVID-19 vaccine. 2021 14 December 2021 16 December 2021]; Available from: https://www.fda.gov/emergency-preparedness-and-response/coronavirus-disease-2019-covid-19/comirnaty-and-pfizer-biontech-covid-19-vaccine.

5. Moderna COVID-19 Vaccine. 2021 13 December 2021 16 December 2021]; Available from: https://www.fda.gov/emergency-preparedness-and-response/coronavirus-disease-2019-covid-19/moderna-covid-19-vaccine.

6. Sadoff, J., et al., Safety and Efficacy of Single-Dose Ad26.COV2.S Vaccine against Covid-19. N Engl J Med, 2021. 384(23): p. 2187–2201.

7. Trinite, B., et al., SARS-CoV-2 infection elicits a rapid neutralizing antibody response that correlates with disease severity. Sci Rep, 2021. 11(1): p. 2608.

8. Carrillo, J., et al., Humoral immune responses and neutralizing antibodies against SARS-CoV-2; implications in pathogenesis and protective immunity. Biochem Biophys Res Commun, 2021. 538: p. 187–191.

9. Garritsen, A., et al., Two-tiered SARS-CoV-2 seroconversion screening in the Netherlands and stability of nucleocapsid, spike protein domain 1 and neutralizing antibodies. Infect Dis (Lond), 2021. 53(7): p. 498–512.

10. Wu, Y., et al., A noncompeting pair of human neutralizing antibodies block COVID-19 virus binding to its receptor ACE2. Science, 2020. 368(6496): p. 1274–1278.

11. Zost, S.J., et al., Potently neutralizing and protective human antibodies against SARS-CoV-2. Nature, 2020. 584(7821): p. 443–449.

12. Piechotta, V., et al., Convalescent plasma or hyperimmune immunoglobulin for people with COVID-19: a living systematic review. Cochrane Database Syst Rev, 2021. 5: p. CD013600.

13. Vandeberg, P., et al., Production of anti-SARS-CoV-2 hyperimmune globulin from convalescent plasma. Tranfusion, 2021: p. 1–5.

14. Fiedler, S., et al., Antibody Affinity Governs the Inhibition of SARS-CoV-2 Spike/ACE2 Binding in Patient Serum. ACS Infect Dis, 2021. 7(8): p. 2362–2369.

15. Bubonja-Sonje, M., et al., Diagnostic accuracy of three SARS-CoV2 antibody detection assays, neutralizing effect and longevity of serum antibodies. J Virol Methods, 2021. 293: p. 114173.

16. Belda, F., et al., Seroconversion panels demonstrate anti-SARS-CoV-2 antibody development after administration of the mRNA-1273 vaccine. medRxiv, 2021.

17. Connor, R.I., et al., Vpr is required for efficient replication of human immunodeficiency virus type-1 in mononuclear phagocytes. Virology, 1995. 206(2): p. 935–44.

18. Sanchez-Palomino, S., et al., A cell-to-cell HIV transfer assay identifies humoral responses with broad neutralization activity. Vaccine, 2011. 29(32): p. 5250–9.

19. Qian, Z., et al., Identification of the Receptor-Binding Domain of the Spike Glycoprotein of Human Betacoronavirus HKU1. J Virol, 2015. 89(17): p. 8816–27.

20. Ou, X., et al., Characterization of spike glycoprotein of SARS-CoV-2 on virus entry and its immune cross-reactivity with SARS-CoV. Nat Commun, 2020. 11(1): p. 1620.

21. Mendrone-Junior, A., et al., Correlation between SARS-COV-2 antibody screening by immunoassay and neutralizing antibody testing. Transfusion, 2021. 61(4): p. 1181–1190.

22. Tan, C.W., et al., A SARS-CoV-2 surrogate virus neutralization test based on antibody-mediated blockage of ACE2-spike protein-protein interaction. Nat Biotechnol, 2020. 38(9): p. 1073–1078.

